# International comparison of the impact of the pandemic and vaccination measures adopted on children and adolescent population

**DOI:** 10.1101/2022.03.24.22272863

**Authors:** Luis Rajmil, María-Camila Pinzón-Segura, Seth Christopher Yaw Appiah, Bernadine Ekpenyong, Fernando González, Zuhal Gündoğdu, Selim Oncel, Özlem Çakıcı

**Affiliations:** Pediatrician and Epidemiology and Public Health Specialist.; Pediatrician, PhD(c) in Public Health at the National University of Colombia; Clinical Epidemiologist Specialist; Medical Sociologist and Infectious disease public health scientist. Department of Sociology and Social Work, Kwame Nkrumah University of Science and Technology,Ghana; Department of Public Health, College of Medical Sciences, University of Calabar, Nigeria; UNICEF Chile Country Office; Kocaeli University, Faculty of Medicine, Head of Social Pediatrics, Kocaeli, Turkey; Kocaeli University, Faculty of Medicine, Head of Pediatric Infectious Diseases, Kocaeli, Turkey; Kocaeli University, Faculty of Medicine, Pediatric Infectious Diseases, Kocaeli, Turkey

**Keywords:** child health, children, COVID-19, adolescents, social inequalities, vaccine

## Abstract

**Background:** The objectives of this study were to compare the cumulative incidence, hospitalizations and mortality, by country and age group, in child and young people (CYP) from the beginning of the pandemic to January 2022, and to describe the differences and similarities and justification in the measures adopted in relation to CYP vaccination against SARS-CoV-2.

**Methods:** A descriptive quantitative summary of the available data on the impact of the COVID-19 on children and adolescents (<18y) from 7 countries (Chile, Colombia, Paraguay, Nigeria, Ghana, Spain) or regions (Kocaeli, Turkey), based on official published data was performed. Outcome measures were: available data on incidence, admission to hospital and to intensive care units (ICU), and deaths, by country or region; vaccination plans, including age, type of vaccine and official justification about the proposal.

**Results:** All countries analyzed data showed variable incidence rate, and relatively low ICU hospitalizations and deaths. Vaccines used and age at starting were also variable, i.e. starting at 3-5y in some Latin American countries, while in other countries proposal starts at 15y old. Almost all justifications were based on the idea to promote collective protection, and that vaccination is important as it -directly and indirectly-protect the rest of the population.

**Conclusions:** The results reinforce the idea of the urgent need to prioritize globally and equitably distribution of vaccines in the population at greatest risk, and to apply the precautionary principle in CYP before deciding to massively vaccinate it.

**What is known:** The child population has less severe infections and seems to transmit the infection less than the adult population.

Published data from the Randomized Clinical Trials (RCT) of vaccines in minors presented short follow-up than in the adult population and basically studied the incidence of infection as the main endpoint

**What this study adds:** Mortality in the child population due to COVID-19 since the start of the pandemic has been low in all the countries analyzed

The vaccines used as well as the vaccination guidelines shows high variability in these countries

The results of the study reinforce the need to establish global and equitable mechanisms for the distribution of vaccines and prioritize the population at greatest risk, such as older people.

In the child population, it is necessary to apply the precautionary principle before establishing massive vaccination plans

## Introduction

Vaccines represent a breakthrough in controlling for and against the spread of many contagious diseases. However, not all vaccines are the same in terms of their origin, mechanism of action, effectiveness and duration,.

COVID-19 vaccines, despite their apparent impact on reducing mortality in the general population and especially in older people, have been approved in an emergency situation and based on phase 3 clinical trials.

The pandemic has lasted almost 2 years, and inequities in the production, distribution and administration of vaccines remain. If the vaccine were the definitive solution for this process, the necessary measures are not being carried out so that the world’s adult population can be immunized, and thus prevent the appearance and spread of new strains.

Vaccines in the child and adolescent population have begun to be tested and administered subsequently to the adult population, which means that there has been some data on evolution for even less time. The secondary effects, and likewise the rest of the intermediate and final results still have some gaps.^1^

The child population so far is the least affected by the pandemic in terms of severity (ICU admissions) and mortality.^2^ There is some certainty that minors are contagious and can infect the rest of the population, but in general they are not usually carriers of significant viral loads. ^3^

Faced with this situation, countries have adopted variable attitudes and measures in relation to immunization of children. Just some countries in Europe and the United States began vaccination with 2 doses of vaccine between 12-15 years old 3-4 weeks apart, and later approved and began vaccination for children under 5-11 years. In the United Kingdom at the moment, 2 doses 8 weeks appart had been proposed for children 12-14y at high risk, and offers a second dose 12 weeks apart for healthy 12-15 years who decide to be vaccinated. ^4^

There are doubts regarding the lack of transparency on the validity of the safety data and side effects both from clinical trials and from monitoring after administration of the vaccine in the general population. ^5^ For this reason –as well as to avoid continuing with what some authors have called “childism”,^6^ which denies giving prominence to minors and creates prejudices against them-, the precautionary principle should be applied with regard to childhood vaccination and wait for a sufficient and necessary observation time to improve the level of evidence of the measures to be adopted.

For this reason, it was proposed to try to answer the following questions: Is it possible to compare the cumulative incidence, hospitalizations and mortality by country and by age group in the child and adolescent population from the beginning of the pandemic to the last available data?. What differences and similarities are in the measures adopted by different countries in relation to childhood and adolescent vaccination against SARS-CoV-2? And what are the justifications related to the use / non-use of these vaccines in children and adolescents in the countries analyzed?

## Methods

A descriptive quantitative summary of the available data from the beginning of the pandemic until January 2022 on CYP SARS-CoV-2 incidence, admissions to intensive care units (ICU), and deaths have been analyzed by age groups, and country or region.

In the same way, the measures adopted regarding the administration of COVID-19 vaccines by age groups of children, the dates on which the vaccination started to be administered, the inclusion / exclusion criteria, availability and type of vaccines, was described.

Finally, a description of official justifications of each country regarding the decisions to vaccinate or exclude children and adolescents was carried out.

Countries included in the study were based on the opportunity and availability of researchers from the International Society of Social Pediatrics, ISSOP (www.issop.org), and the International Network for Research on Inequalities in Child Health (INRICH, https://inrichnetwork.org/) COVID-19 working group. This group started working at the beginning of the pandemic analysing the impact of the pandemic on child health. Finally, data from seven countries (Chile, Colombia, and Paraguay from Latin American countries, Spain from Europe, Ghana and Nigeria from Africa, and the region of Kocaeli from Turkey, the latest based on the Kocaeli University Hospital data) contributed to the analysis of the overall situation in the current work, although it would not represent the situation at the whole group of countries/participants.

The source of the data analyzed were the official websites and/or official publications of each participant country (i.e. Governments or Ministries of Health, etc.) or data extracted from publications in indexed journals. For example, in the case of Spain the data collected was based on the latest published information on the evolution of COVID-19 pandemic reported by the Spanish Ministry of Health (updated on December the 9^th^, 2021: “*Situación de COVID-19 en España a 9 de diciembre de 2021. Equipo COVID-19. RENAVE. CNE. CNM (ISCIII)”*, the updated report on hospitalization and death,^7^ and the available data from the Spanish National Institute of Statistics (https://www.ine.es/jaxiT3/Datos.htm?t=31304); the latest was collected to estimate indicators analyzed at the Spanish general population level 0-19y.

## Results

### Incidence, hospitalization and mortality

Cumulative incidence and hospitalizations have been presented in the countries analyzed, through available data and showed a high variability. Mortality due to SARS-CoV-2 from the start of the pandemic showed a relatively low number of cases in the CYP in all the countries analyzed (n= 127 in Chile, n= 263 -1.8/100000 h. in Colombia, n= 49 in Paraguay, n= 4 in Ghana, n= 26 in Nigeria, n= 40 - 0.43/100000 h. in Spain, and 3 deaths in Kocaeli (table 1).

**Table 1.**
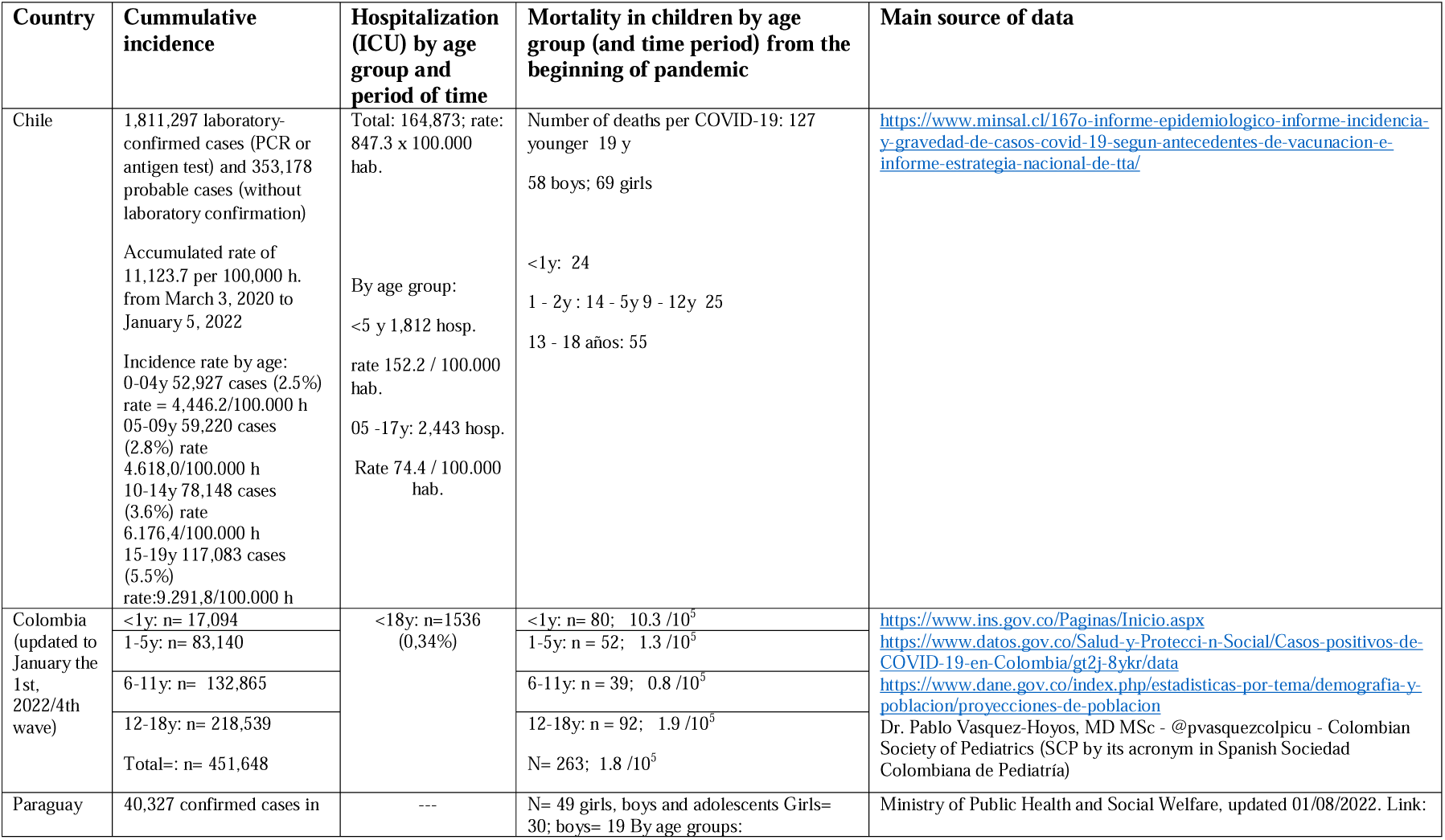
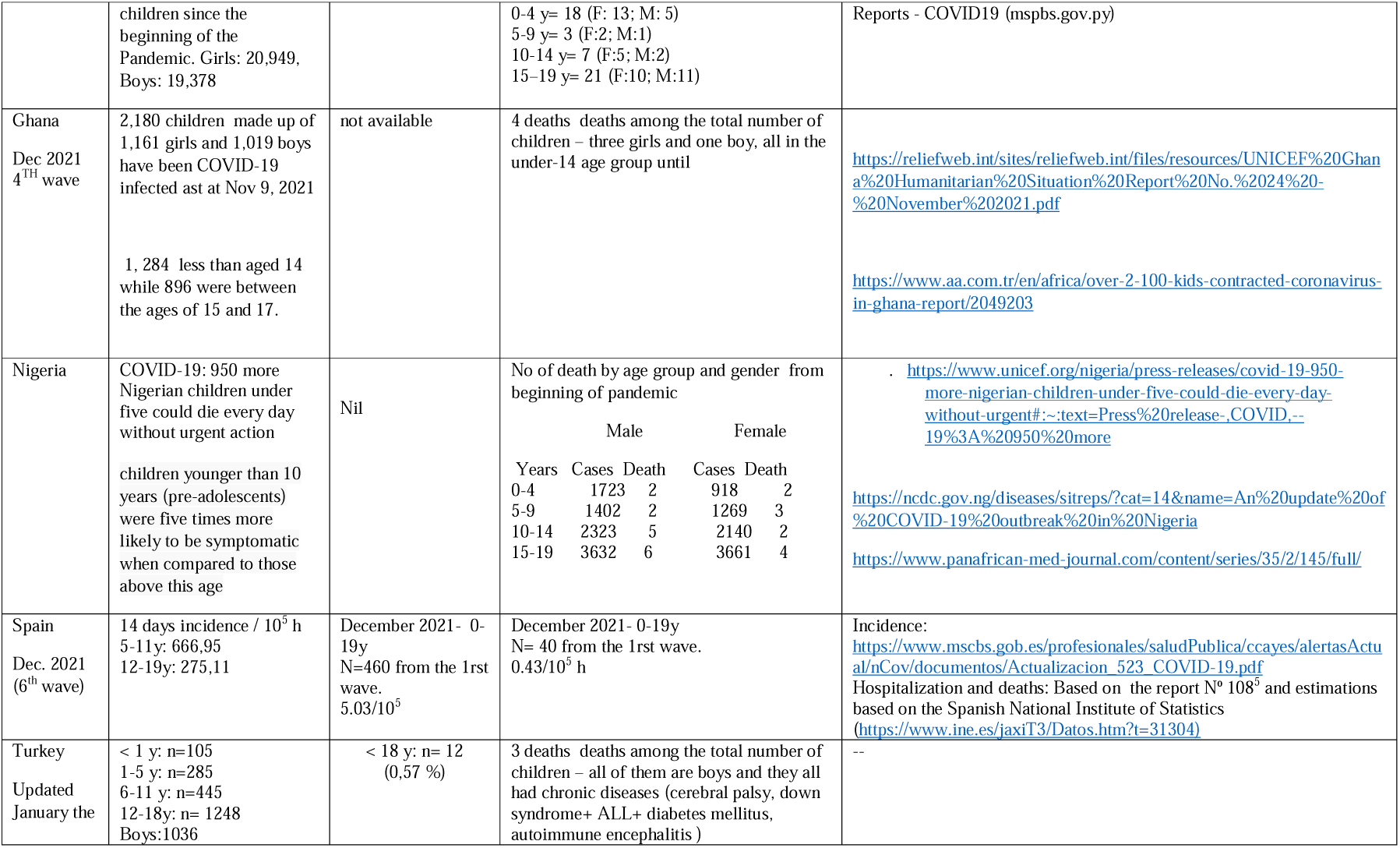
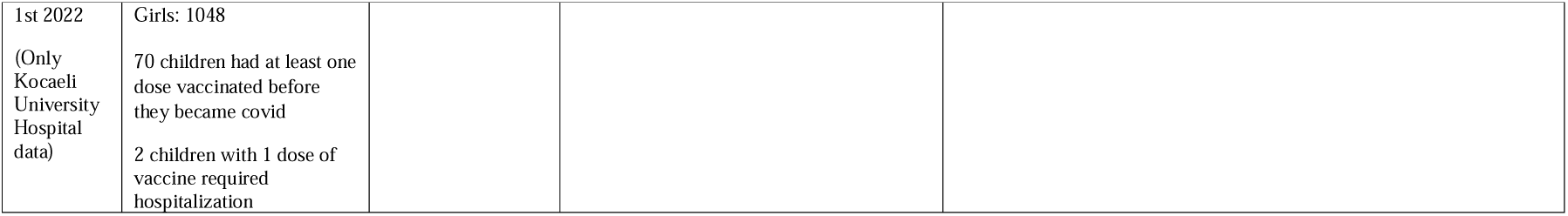
Incidence, hospitalization (intensive care units, ICU), and mortality due to COVID-19.

### Vaccinating children and adolescents and justification

The start of the vaccination plan and the type of vaccine in CYP show differences according to the countries analyzed (Table 2). While in Latin American countries, the vaccination plan has been established throughout the second half of 2021, from age of 3 (in Chile, Colombia) and 5 (in Paraguay) years old, in Ghana it is proposed to vaccinate from 15 years onward, beginning in January of 2022. Spain, for its part, shows CYP vaccine plans beginning in July 2021 for CYP older than 11y and starting in December of 2021, with children who are 5 years old or more. Sinovac was the vaccine chosen for children under 12 in Colombia and Chile, while Sinovac or Pfizer in Paraguay. In Colombia and Paraguay all children above 11 are vaccinated with Pfizer, while Chile has implemented Pfizer or Sinovac for the same range of age. To date, in the European Union, the only vaccine approved in children younger than 12y is Pfizer, while in Turkey started with Sinovac and Pfizer.

**Table 2.** Age at starting vaccination, and type of vaccine.

| Country | Vaccines included |  |  |  |
| --- | --- | --- | --- | --- |
|  | Age group | Proposed vaccine, number of doses, and interval | Data starting vaccination | Observations |
| Chile | 3-5 y | Coronavac 2 dosis, 28 days | 6 december 2021 | With the authorization of parents |
|  | 6-11 y | Coronavac 2 dosis, 28 days | 13 september 2021 | With the authorization of parents |
|  | 12-18 y | Pfizer 2 dosis, 28 days | 21 june 2021 | With the authorization of parents |
|  | 12-18 y | Coronavac 2 dosis, 28 days | 13 september 2021 | With the authorization of parents |
| Colombia | 3-11y | Sinovac Biotech vaccine (inactive virus), two doses 4 weeks apart | 31/10/21 | Based on a two phases strategy, Colombia Health Ministry decreed 3 steps in the first phase, looking to reduce COVID severe morbidity and mortality and 2 steps in the second one to reduce the contagiousness. Two of the main supportive documents of this strategy were: 1) 466 Decree, May 8th, 2021; and 2) General Guideline for the expanded program of immunizations (PAI by its acronym in Spanish Plan Ampliado de Inmunización) in the COVID pandemic context - Jun, 2021).<br>Despite Colombian government has emphasized the importance of "presence educational methodology" what has happened is that the CYP isolation was one of the first measures taken on practice<br>Vaccination in this population will always be given in the presence of the parents or legal guardian of the child/young, and the informed consent must be signed by both of them.<br>The Scientific Committee of the Colombian Ministry of Health and Social Protection, supported by the Colombian Society of Pediatrics (SCP by its acronym in Spanish Sociedad Colombiana de Pediatría), the Colombian Association of Infectology, the Colombian Association of Immunology, the Colombian Institute for Drug and Food Surveillance (INVIMA by its acronym in Spanish Instituto Nacional de Vigilancia de Medicamentos y |
|  | 12-14y (without comorbidities) | Pfizer-BioNTech vaccine (ARNm), two doses 3-4 weeks apart | 28/08/21 |  |
|  | 15-17y (without comorbidities) |  | 20/08/21 |  |
|  | 12-17y (with comorbidities) |  | Jun/2021 |  |
|  |  |  |  | Alimentos), and the current Colombian regulation (referred above) approved the use of new vaccines in the pediatric population<br>In any case, CYP with a health problem or disease in treatment (previously not classified as comorbid by COVID guidelines) must attach a medical concept to a better guide of the process and define whether there is or not a specific restriction on immunization |
| Paraguay | 5-11y | Coronavac and Pfizer | 01/17/2022 (subject to confirmation) | As a strategy to facilitate the vaccination of adolescents, the Ministry of Public Health and Social Welfare established "a special day", in which adolescents could preferentially go for vaccination. |
|  | 12-18y | Pfizer | 07/23/2021. Adolescents with underline pathology<br>10/13/2021. Adolescents without pathology |  |
| Ghana | 15 years above | No new proposals on how the vaccine is to be taken by children . | January 2022 | All children 15 years plus in secondary and tertiary schools |
| Nigeria | The United Nations Children's Fund (UNICEF), partnering the Federal Government, will soon commence immunisation of children in Nigeria against the Coronavirus Disease (COVID-19). |  |  | https://guardian.ng/news/unicef-partners-fg-on-childrens-covid-19-vaccination/ |
|  | To tackle these outbreaks, the federal government of Nigeria, through the National Primary Health Care Development Agency (NPHCDA), has adopted the “whole family” approach in the second phase of COVID-19 vaccination.<br>The “whole family” approach combines COVID-19 vaccination with healthcare services like childhood vaccination, malnutrition and screening for non-communicable diseases. |  |  | https://www.gavi.org/vaccineswork/combining-covid-19-and-routine-vaccination-nigeria-implements-whole-family-approach |
| Spain | 5-11y | ARNm, 2 dosis (10 mcrg, Pfizer vaccine), 4-8 weeks apart | 13/ Dec / 2021 | All childhood population included without exceptions |
|  | 12-18y | ARNm (30 mcrg, Pfizer vaccine) 3-4 weeks apart | July /2021 | All adolescent population included without exceptions |
| Turkey | 15-18 y ( with or without comorbidities)<br>12-15 y ( with comorbidities) | Sinovac Biotech vaccine (inactive virus), two doses 4 weeks apart<br>Pfizer-BioNTech vaccine (ARNm), two doses 3-4 weeks apart<br>3 months after 2nd dose, 3rd dose recommended for both vaccines | 16/08/2021 | Individually determined by the national information system |
|  | 12-15 y ( without comorbidities) | Sinovac Biotech vaccine (inactive virus), two doses 4 weeks apart<br>Pfizer-BioNTech vaccine (ARNm), two doses 3-4 | 05/09/2021 |  |
|  |  | weeks apart<br>3 months after 2nd dose, 3rd<br>dose recommended for both<br>vaccines |  |  |

All countries have as justification for the CYP vaccination program, its direct and indirect potential benefits (herd effect) transmitting the idea that “vaccination protects against virus transmission” (see Supplementary material). Some other claims to promote vaccination were: scientific evidence which support that the immunization benefits are greater than its risks; the need to return to normality, with a special regard to reactivate the economy; and reduce the specific mortality and incidence of severe cases, particularly related with new variants.

## Discussion

The results confirm that, in the countries analyzed, and compared to the general population, the incidence of severe disease and mortality in the CYP due to SARS-CoV-2 was low. The work also reflects the variability in the vaccination schedule and that the main justification for starting the vaccines in the CYP was the protection of the general population and the impact of COVID-19 on social and economic activities more than the impact on child health. There are a few clinical outcome studies, with CYP participation, that lead to the decsision to vaccinate CYP despite its low SARS-CoV-2 mortalities.

This work highlights the urgent need to carry out global approaches to CYP vaccination based on equity and its essential characteristics and needs. As happened at the beginning of the pandemic, the child population was stigmatized and suffered the adverse consequences of the measures taken to mitigate the pandemic.^8^ The lack of inclusion as social and legal subjects has also been shown.^9 10^ Similarly, the inclusion of the child population in vaccination campaigns has been based more on the protection of the general population than on their own benefit.

It must be taken into account that the COVID-19 pandemic has exacerbated pre-existing social inequalities. Excess mortality both within and between countries, at higher income-level as well as low and middle income countries, has occurred in the population at greater risk of poverty and marginality conditions, which especially affects the child population. ^11^ Moreover, in some countries such as Nigeria, it was estimated an excess of mortality of 950 more children under five every day at the beginning of the pandemic associated to the COVID-19 pandemic disruption of routine services and threatens to weaken the health system.^12^ On the other hand, study from 6 sub-Saharan African countries showed that morbidity and mortality were substantially higher than reported among those in non-African settings, mainly associated with noncommunicable disease comorbidities. ^2^ In Colombia, since the beginning of the pandemic the rates of violence against children, especially in the domestic scenario, have significantly increased, data reproduced in other latitudes as it was informed by WHO. ^13^

Another aspect that draws attention is the type of vaccine approved and marketed in the participating countries, which probably reflects political and economic decisions that have nothing to do with scientific evidence. ^14^

As has been stated, childhood vaccines have been approved in an emergency, and with a very short follow-up. Expedited approvals are associated to post-authorisation studies to confirm that the medicines safely, but a long history of concerns has emerged about given that post-authorisation studies often fail to deliver, many take years longer than planned, and so on.^15^ Among the results analyzed in the clinical trials, mortality, severity, and the level of transmission and viral load in the vaccinated have not been taken into account as end points. ^16 17 18^ For this reason, the justification that has been expressed to promote vaccination in adolescents, especially in children under 11y is striking.

Moreover, adult immunisation is feasible with the right investments and actively pursued in almost all countries; a situation that is different in CYP, since the immunization benefits in this population (primary to reduce the risk of severe disease and death) are much less strong than those associated with vaccinating older adults. The benefits of vaccinating children to reduce the risk of severe disease and death are much less than those associated with vaccinating older adult. ^19^ In summary, the equitable distribution of vaccines in the population at greatest risk should be prioritized and its distribution worldwide before proposing the vaccination of CYP. ^20^

Among the limitations of the study, particularly its lack of global representativity related with a convenience selection of the participant countries, it is worth mentioning that the fast and variable Sars-CoV-2 pandemic dynamics press to have variable and quick measures; the epidemiologic definitions have changed over time, as well as data collection, the number and specificity of tests that are carried out, and other variables.

After the caveat referred above, the results of the research, in terms of severity and mortality, confirm that regardless of the level of development and access to vaccines, the SARS-CoV-2 affections to the CYP are similar in the countries analyzed, confirming that this population presents fewer negative results compared to the rest of age range populations.

As a summary, the results of the present study reinforce the idea of the urgent need to prioritize globally and equitably vaccine distribution in the population at greatest risk, and to apply the precautionary principle in the CYP before deciding to massively vaccinate it.

## Data Availability

All data are available at the links provided in Tables 1 and 2 of the manuscript and/or at the Supplementary material

https://www.minsal.cl/167o-informe-epidemiologico-informe-incidencia-y-gravedad-de-casos-covid-19-segun-antecedentes-de-vacunacion-e-informe-estrategia-nacional-de-tta/

https://www.ins.gov.co/Paginas/Inicio.aspx

https://www.datos.gov.co/Salud-y-Protecci-n-Social/Casos-positivos-de-COVID-19-en-Colombia/gt2j-8ykr/data

https://www.dane.gov.co/index.php/estadisticas-por-tema/demografia-y-poblacion/proyecciones-de-poblacion

https://reliefweb.int/sites/reliefweb.int/files/resources/UNICEF%20Ghana%20Humanitarian%20Situation%20Report%20No.%2024%20-%20November%202021.pdf

## Authors contribution

LR conceptualised the paper, and wrote the initial draft. LR, MCPS, SCYA, BE, FG, ZG, SO, and ÖÇ contributed to search and analyze the data. SCYA and MCPS revise the manuscript. All authors approved the final version of the manuscript.

## Patient consent for publication

Not required.

## Ethical approval statement

The study does not require ethical approval. It include data involving information freely available in the public domain (e.g. published and freely available data), and these data were properly anonymised.

