## Supplemental Table 1 for "International comparison of the impact of the pandemic and vaccination measures adopted on children and adolescent population"

### Supplemental material. Summary of main justification on how they propose/recommend vaccinating child and adolescent poulation

| Country | Dates | Main justifications | Source of data |
|---|---|---|---|
| Chile | 2 dec 2021<br><br><br><br><br>27 sept, 2021<br><br><br><br>22 june, 2021 | "The vaccine is voluntary, but we all have a moral obligation to get vaccinated because not only do we protect our health, if we do not get vaccinated we are putting the health of the people we love most, our families, our loved ones, our co-workers, at risk. our community"<br><br>"This is great news for school-age children and for those minors who were not yet considered in the vaccination plans, so they can return to school safely," | https://www.minsal.cl/covid-19-vacunacion-para-ninos-de-3-a-5-anos-parte-el-proximo-lunes/<br><br>https://www.minsal.cl/autoridades-dan-inicio-a-vacunacion-escolar-contra-covid-19-en-ninos-de-6-a-11-anos/<br><br>https://www.minsal.cl/presidente-pinera-da-inicio-al-proceso-de-vacunacion-contra-el-covid-19-para-menores-de-18-anos-la-vacuna-es-voluntaria-pero-todos-tenemos-una-obligacion-moral-de-vacunarnos/ |
| Colombia | | 1. COVID infection and its complications has been increasing<br>The pandemic began to affect the CYP. Although the incidence of infection in those is low, it has been increasing, as well as the cases of severe symptomatology and death attributed to the virus. Indicators in this population (rate of complications, for example) are higher than other diseases for which they are routinely vaccinated, for example Hepatitis A and Influenza.<br><br>It is particularly noteworthy the evidence -socialized by the WHO- regarding the development of multisystemic inflammatory syndrome (SIM) related (chronologically) with the presence of SARS-CoV-2 in CYP.<br><br>2. Based on scientific *evidence the immunization benefits are greater than its risks*<br>It was shown by scientific evidence that the vaccine is safe, well tolerated and produces adequate immunogenicity. The benefits (lower disease burden, lower hospitalization rate, and lower probability of death attributed to infection) are greater than the risks (which are mainly mild adverse effects, near the time of application, as redness or swelling, tiredness, muscle pain, fever or nausea).<br><br>The likelihood of long-term effects such as diabetes or myocarditis is low, but not entirely ruled out, as there is still not enough accumulated monitoring and follow-up time to confirm this suspicion. Some studies showed that RNAm vaccines, such | https://consultorsalud.com/bogota-ya-vacuna-covid-ninos-3-anos/<br><br>https://www.eltiempo.com/salud/vacunacion-covid-19-en-ninos-en-colombia-lo-que-debe-saber-629468<br><br>https://www.javeriana.edu.co/pesquisa/vacuna-covid-19-para-ninos-y-ninas-menores-de-12-anos-esta-todo-claro/<br><br>https://www.minsalud.gov.co/Paginas/Llego-el-turno-de-vacunar-a-menores-entre-los-12-y-14-anos-.aspx<br><br>https://www.minsalud.gov.co/Paginas/Avanza-la-vacunacion-en-ninos-de-3-11-anos.aspx<br><br>https://www.minsalud.gov.co/Paginas/Sociedades-cientificas-respaldan-vacunacion-en-ninos-de-3-a-11-anos-.aspx<br><br>https://www.facebook.com/MinSaludCol/videos/744272476968377 |

| | | | |
|---|---|---|---|
| | | as Pfizer-BioNTech and Moderna, could produce myocarditis in a minimal percentage; however, this disease would be transient and without lethal outcomes.<br><br>Is taken as reference scientific studies from countries where vaccination had begun in CYP; mainly China (security and effectiveness) and Chile (security).<br>References is also made to the following studies:<br>- The LANCET (December 2021) where it was demonstrated than CoronaVac (SINOVAC) was well tolerated and safe, and induced humoral responses in CYP between 3 to 17 years. Titres of neutralizing antibodies induced by the dose of 3.0µg were higher than those of the dose of 1.5µg. The results support the use of the 3.0µg dose with a schedule of two immunizations for studies in CYP.<br>- NEJM (New England Journal of Medicine): Pfizer vaccine is completely safe in children between 12 and 15 years old, produced a greater immune response than in young adults and was highly effective against COVID-19<br>- FDA (Food and Drug Administration Agency): reviewed a study of more than 2,200 American children between the ages of 12 and 15. A week after the administration of the second dose, the research showed no case of COVID-19 in the 1005 children who received the Pfizer-BioNTech vaccine. Among the 978 children who received placebo, there were 16 cases of COVID-19. None of the children were previously diagnosed with COVID-19. The results suggest that the vaccine is 100% effective in preventing COVID-19 in this age group.<br><br>3. Collective protection (Herd effect)<br>Reduce the risk of infection in other populations. Children and young persons are vectors (transmitters) of the virus and may be spreading the disease in homes and communities. Individual protection, but above all collective protection.<br><br>4. Financial stabilization and economic upturn<br>Need to return to presence activies in safe environments, such as educational institutions. Need for economic, social and cultural resumption.<br><br>5. Control of COVID-19 spread and its new variants<br>Reduce the specific mortality and incidence of severe cases, particularly in new variants with higher transmissibility rates (e.g., delta, omicron). The pandemic continues. Prevention. Other peaks are coming | https://www.facebook.com/SociedadColombianadePediatria/videos/vacunación-en-tiempos-de-covid-19-un-reto-para-todos/601096590617209/<br><br>https://www.facebook.com/SociedadColombianadePediatria/videos/foro-vacuna-covid-en-mayores-de-6-años/589894605556607/ |

| | | | |
|---|---|---|---|
| | | (In connetion with the first justification) The characteristics of the pandemic allow to predict that it will continue, as new variants of the virus and outbreaks will come. As populations progressively have been vaccinated, especially adult one, and the virus continues to circulate, CYP are becoming more susceptible to infection.<br><br>6. Children´s Rights approach<br>Children and young persons must be taken into account in vaccination plans, since they were active members of the society to which they belonged. If compliance with the vaccination scheme against COVID is a condition of accessibility or restriction to community-specific activities, the non-inclusion of CYP may limit their participation in certain spaces and events. It is important that they can benefit from this right.<br>7. Ensure compliance with the implementation of COVID public policies<br>In accordance with the vaccination strategy against COVID (normative decree/guideline), which prioritized, by stages based on risk groups, Colombia had cheked with the conditions to begin with this age group, after a progressive and programmed plan that advanced in other age groups | |
| Paraguay | | Ministry of Health: "it is a great satisfaction to be fulfilling this pending debt, the children deserve this vaccination."<br>"It is very important that our children get vaccinated because it has been shown that not only is the acute condition increasing in children, but that it involves absenteeism in some social activities that they deserve to have, and the second component is that it also causes complications, since within four to six weeks the appearance of multisystem inflammatory syndrome is an entity that also requires intensive care and often also a serious evolution; all this can be avoided with a vaccination" | Published in the Paraguayan Information Agency . Statements from the Director of the Expanded Program of Immunizations of the Ministry of Public Health and Social Welfare of Paraguay. January 7, 2020.<br><br>https://www.ip.gov.py/ip/ministerio-de-salud-habilita-inscripcion-para-la-vacunacion-anticovid-de-5-a-11-anos/ |
| Ghana | January 2022 for vaccination to to commence | That the Ghana Public Health Act , ACT 851 of 2012 and its emergency clasues bestows on the Minister of Health to make declarations for which targetted groups should be provided with mandatory Covid-19 vacines. The targetted groups mentioned included staff and students 15 years and above in secondary and tertiray schools . The rational was to prevent further spread of the virus. Children aged 15 years plus form part of the target groups requing attention | Minister of Health, republic of Ghana meet the press<br>https://fb.watch/a0BxyK2LqV/<br><br>Report from the Director General of the Ghana Health Service<br>https://www.reuters.com/world/africa/ghana-make-covid-19-vaccine-mandatory-targeted-groups-january-2021-11-28/ |

| | | | |
|---|---|---|---|
| Spain | Juny 2021 start vaccinating 12-19y | "Vaccination is doubly important as it directly protects each vaccinated person, but it also indirectly protects the rest of the population. The more people are immunized, the less likely it is that the rest (particularly those most vulnerable to severe disease) will be exposed to the virus, or at least to high viral loads."<br>"To protect vulnerable people and the rest of the population" | https://www.mscbs.gob.es/profesionales/saludPublica/prevPromocion/vacunaciones/covid19/docs/Vacuna_COVID_adolescentes_PreguntasyRespuestas.pdf |
| | Nov 2021 Statement for vaccinating children 5-11y | The risk that a child may have complications that require hospitalization for COVID-19 is not absent. Children aged 5-11y accounted for 6.6% of COVID-19 cases during the previous (fifth) wave and 66.2% of cases for children under 12. 0.21% required hospital admission (of which 68% did not have a underlying disease), 0.016% were admitted to the ICU and a child with a severe underlying disease died (0.001%).<br>Children can suffer, a few, but severe disease | Example from Ministry of Health of Catalonia, Spain<br>https://canalsalut.gencat.cat/web/.content/_A-Z/V/vacuna-covid-19/ciutadania/vacunacio-infantil/preguntes-frequents-vacuna-menors.pdf<br>And the Catalan Pediatric Soc.<br>https://www.academia.cat/files/204-9509-FITXER/docscpvacuna511a1.pdf |
| Turkey | | ''It is recommended that all children over the age of 12 in the world and in our country should be vaccinated against COVID-19. ''<br>This is the only suggestion on the official website of the Turkish Ministry of Health ın response to question 58. | **https://covid19asi.saglik.gov.tr/TR-77694/sikca-sorulan-sorular.html?Sayfa=4** |
